# Clinical genetics laboratories use divergent demographic frameworks across countries: comparing data structures for ‘race’, ‘ethnicity’, and ‘ancestry’ on test requisition forms

**DOI:** 10.1101/2021.12.17.21267965

**Authors:** Alice B. Popejoy, Julia Gimbernat Mayol, Katherine Anderson, Gillian Hooker

**Affiliations:** Department of Biomedical Data Science, Stanford University School of Medicine; Department of Public Health Sciences, University of California, Davis; Department of Biomedical Engineering, Imperial College of London; Concert Genetics, Inc

**Keywords:** Race, Ethnicity, Genetic ancestry, Geographic ancestry, Biogeographic ancestry, Biological racism

## Abstract

**Purpose:** The goal of this study is to investigate how population groups are represented on requisition forms for clinical genetic testing in different laboratories.

**Methods:** Clinical laboratory test requisition forms (RFs) were obtained from 70 laboratories in the US, Canada, Europe, and Australia. Details about the laboratories and how RFs represent patient demographics were extracted and analyzed for trends between forms in the U.S. (N=213) and other countries (N=203).

**Results:** Clinical genetics laboratories included in the analysis vary widely regarding the format of demographic data collected on test requisition forms. US-based laboratory RFs are more likely than those from other countries to include ‘race’ or ‘ethnicity’. These are most often represented as categorical data, with multiple-choice options. RFs from laboratories in other countries do not include ‘race’, and those that include ‘ethnicity’ most often provide a blank space for open-ended responses.

**Conclusions:** These results are consistent with existing research on heterogeneity in the nomenclature and number of categories used to describe patient populations across clinical genetics laboratories in the US. It also suggests systemic differences in the way measures of diversity are conceptualized in the US compared to other countries.

## INTRODUCTION

Clinical genetics laboratories provide requisition forms (RFs) for clinicians to complete when ordering tests, and the content of these forms may vary depending on the type of test. Prior research has shown that patient demographics are characterized in different ways on test requisition forms across laboratories in the US.^1^ The purpose of this study is to further investigate the use of population descriptors on RFs in the US, compare these to RFs from clinical genetics laboratories in other countries, and assess trends.

To interpret regional and cultural trends in population descriptors and groupings, here on referred to as *demographic frameworks*, we must first acknowledge that people have varied conceptual understandings of the terminology used to describe human populations.^2,3,4^ These conceptual frameworks are based on cultural context(s),^5,6,7^ and institutions define populations of interest in different ways to achieve various objectives.

This study provides a comparison of demographic frameworks used in clinical genetics laboratories across regional contexts, to illustrate how they may be influenced by historical, social, cultural, and political factors. We offer an interpretation of differences observed between US and non-US laboratories and suggest next steps.

## MATERIALS & METHODS

### Clinical Laboratory Selection

Requisition forms (RFs) from 70 clinical genetics laboratories were collected in June 2018. Laboratories were selected for analysis among those rated as highly active in clinical curation by Concert Genetics, and ranked in ClinVar Miner^8^ based on the number of variants they had submitted to ClinVar.^9^ Starting with the largest number of variants submitted, we searched each laboratory website for blank copies of all test requisition forms. If RFs were not available online for a given laboratory, the next one down on the list was selected until 35 US laboratories and 33 non-US laboratories were identified. All laboratories selected had submitted a minimum of 30 variants to ClinVar at the time of analysis.

The US National Center for Biotechnology Information (NCBI) Genetic Testing Registry^10^ was used to identify 2 additional non-US laboratories in countries not yet represented in the selected group to facilitate a balanced comparison between US-based and international laboratories.

### Requisition Form Processing

All RFs that were available online were downloaded and processed, including translation into English (using Google Translate) for 19 laboratories. Metadata from all RFs were then recorded in an Excel spreadsheet, including: the name of the laboratory and country in which it operates; [if in the U.S.] whether the lab accepts samples from international clients; the title of the RF; clinical specialty to which it pertains; and [if non-US] the language(s) in which it is written. Details about the population framework on each RF were coded by: 1) format of any question related to patient population (i.e., open-ended or multiple-choice); 2) the terminology used to describe such groups (e.g., race, ethnicity, ancestry, nationality, geographic origin, or other related descriptor); 3) specific categories included as multiple-choice options (if applicable); and 4) whether or not free-text entries were permitted in association with an ‘other’ category (if applicable).

### Comparative Analysis of Population Measures

When multiple RFs from the same laboratory used the same population framework, one of those forms was selected to represent all others with the identical demographic format. The following metadata from these unique forms were compared between the US and other countries: 1) the proportion of laboratories that use RFs with a structured population framework; 2) terms and other characteristics of the framework(s) used; and 3) and the co-occurrence of categories across forms using a multiple-choice format. We hypothesized that RFs from US clinical laboratories would use different population frameworks than other countries.

## RESULTS

Requisition forms (RFs) from US clinical genetics laboratories are more likely to include information about patient race, ethnicity, or ancestry (REA) than those in other countries (Fig.1). Among those who do ask about patient REA on RFs, the US is the only country in the analysis whose clinical laboratory RFs include the word ‘race’, whereas laboratories in other countries tend to use ‘ethnicity’, in addition to rare uses of ‘nationality’ and ‘geographic origin’ (Fig. 2). Additionally, U.S. laboratories are more likely than those in other countries to structure the question using multiple-choice categories for REA, and 74% of those include an ‘Other’ category to specify an alternative response (Fig. 3). In contrast, 89% of RFs from non-U.S. laboratories (i.e., European countries, Canada, and Australia) that ask about REA only offer a blank text option for patients or providers to write in open-ended responses.

**Figure 1.**
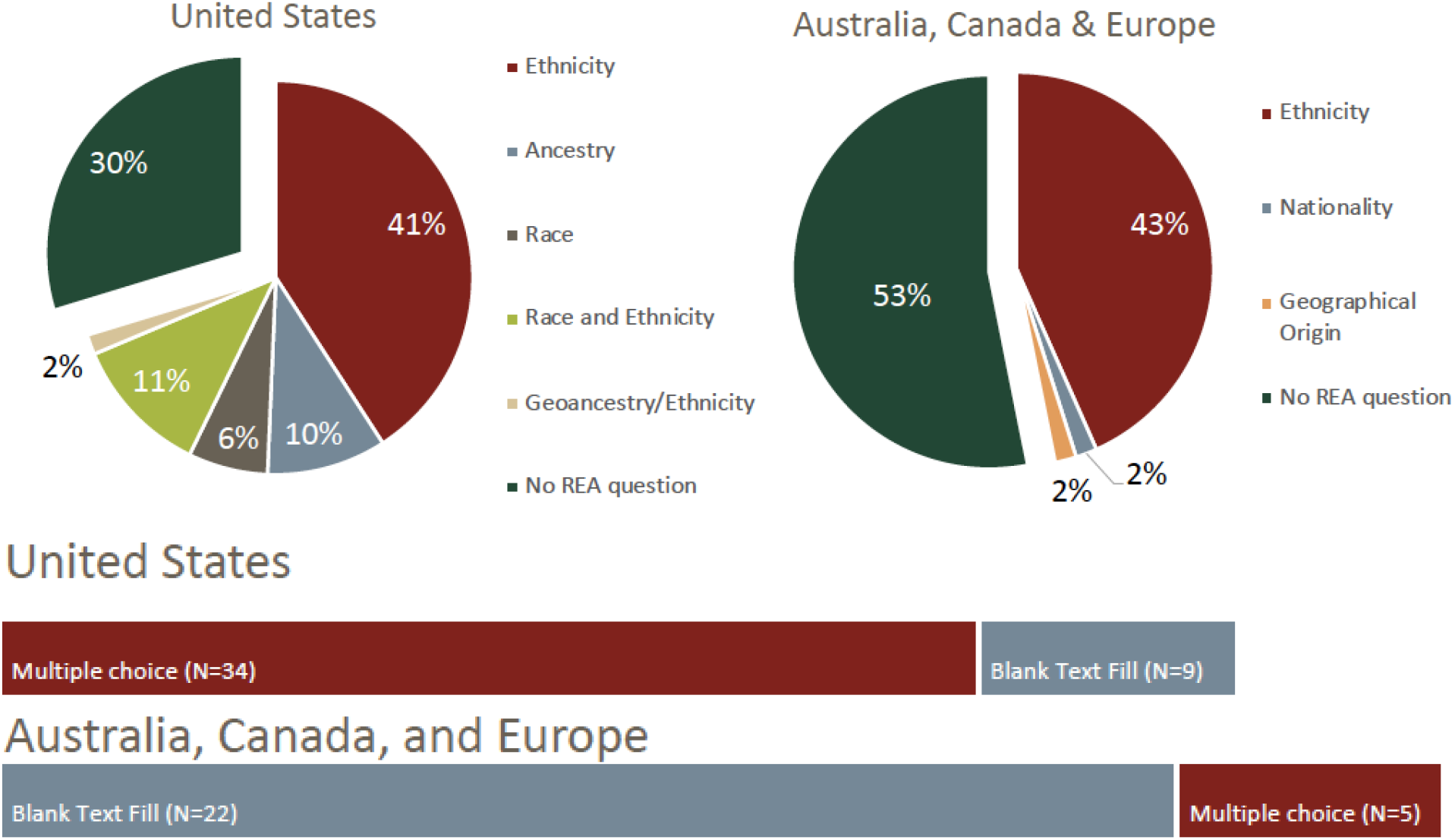
Proportion of US and non-US clinical genetics laboratories that include questions about patient race, ethnicity, ancestry, or other background information.

**Figure 2.**
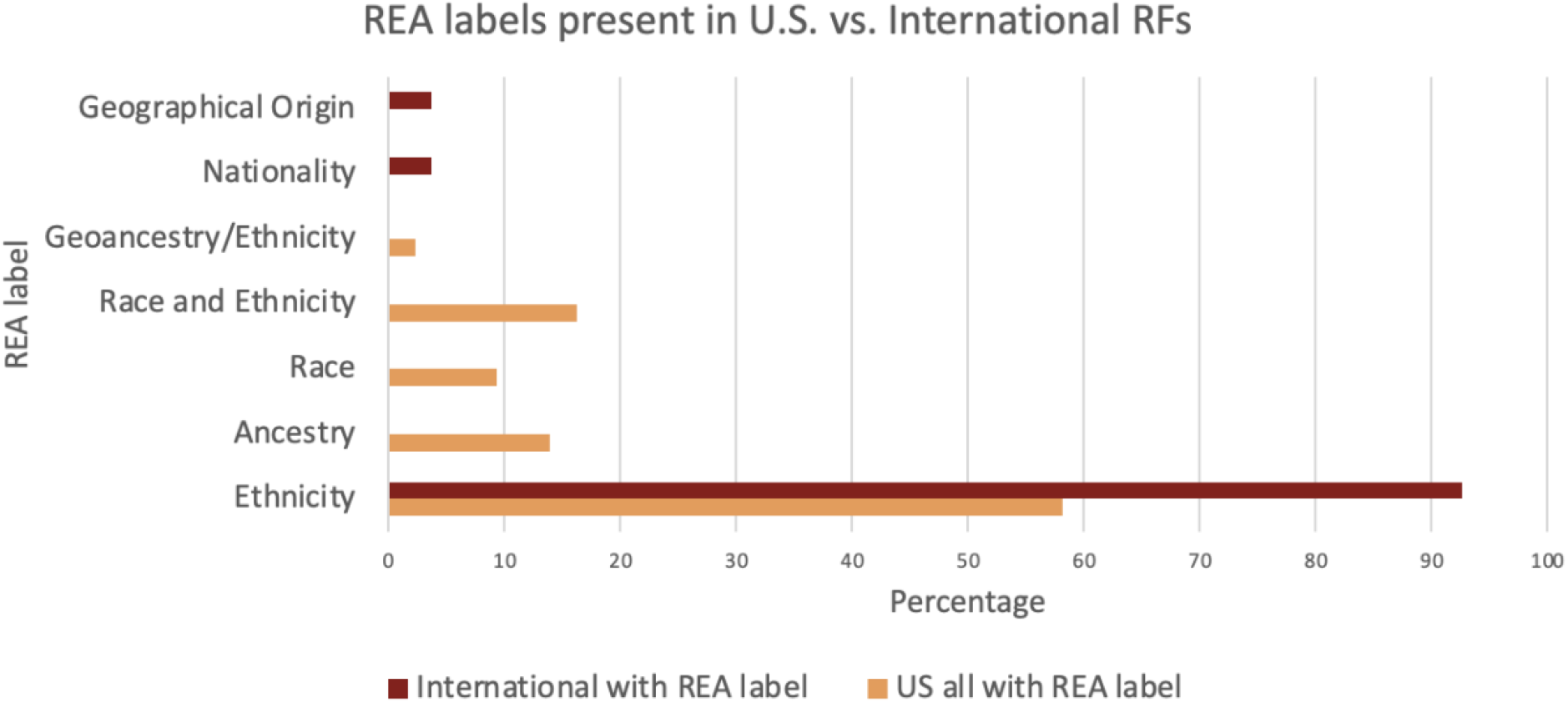
Terminology used to describe populations on RFs from U.S. and non-U.S. labs with questions about patient race, ethnicity, ancestry, or related information (REA).

**Figure 3.**
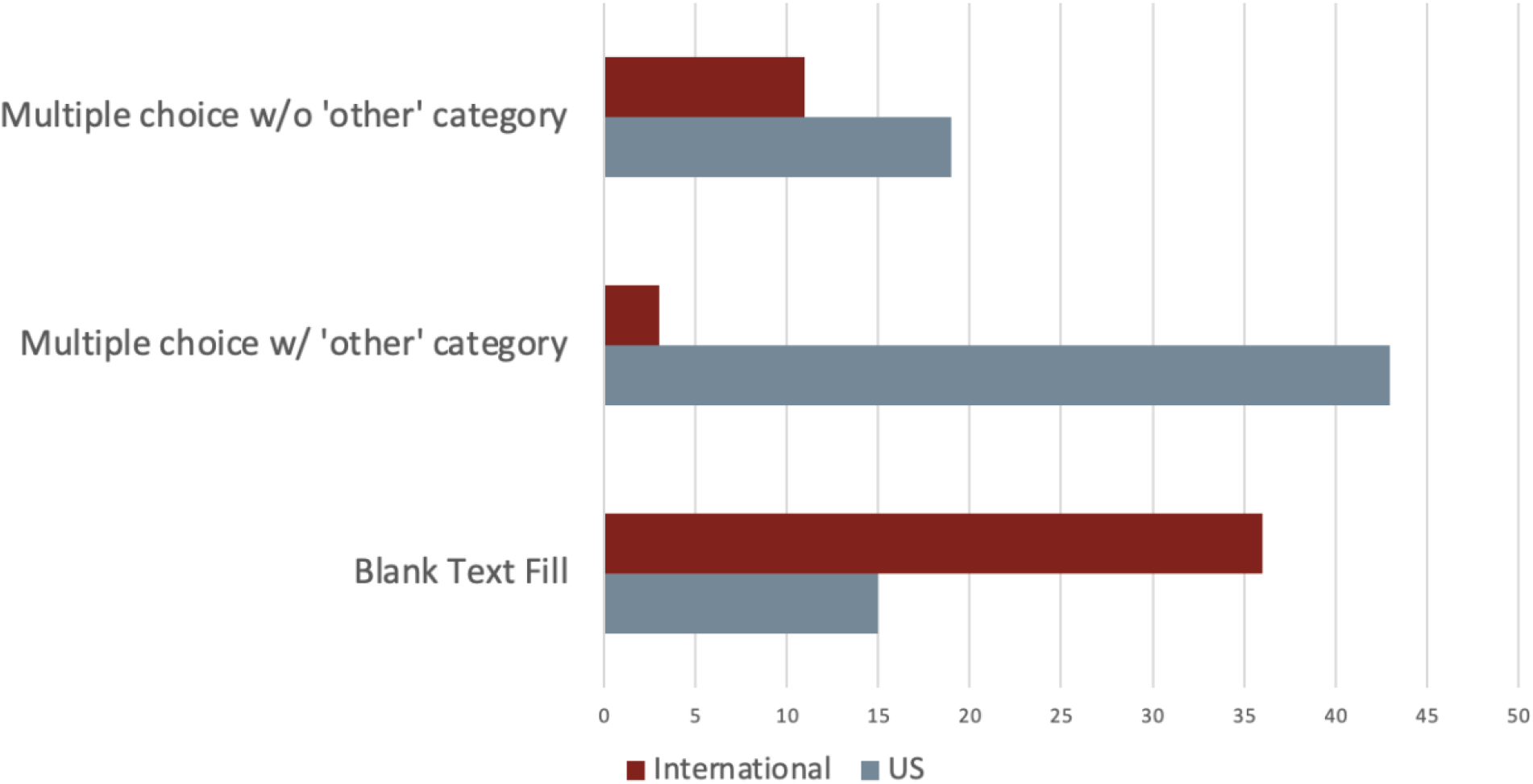
Number of requisition forms from U.S. and non-U.S. (“International”) clinical laboratories that collect information about patient background using multiple choice only, multiple choice with an ‘other’ option to fill in the blank, and blank text only formats.

Of the 35 U.S.-based genetic testing laboratories analyzed, 25 explicitly offer testing to international clients. Most of these laboratories require international ordering providers to use the same RF as their American counterparts, many of which use American-specific terminology in their population frameworks such as “African American” or “Asian American.” Two laboratories offered separate RFs for international samples, however neither of these laboratories removed the American-specific terminology from their forms. Figure 4 illustrates the population descriptors (categories) that are most used on forms from U.S. and non-U.S. labs, and Fig. 5 shows how often these descriptors occur together on the same form. Groups of similar descriptors are often used interchangeably with one another, and among these groups, the most common to co-occur are: 1) ‘Black’, ‘African American’, or ‘African’, 2) white, 3) Asian, and 4) ‘Hispanic’, ‘Latino’, or ‘Spanish’.

**Figure 4.**
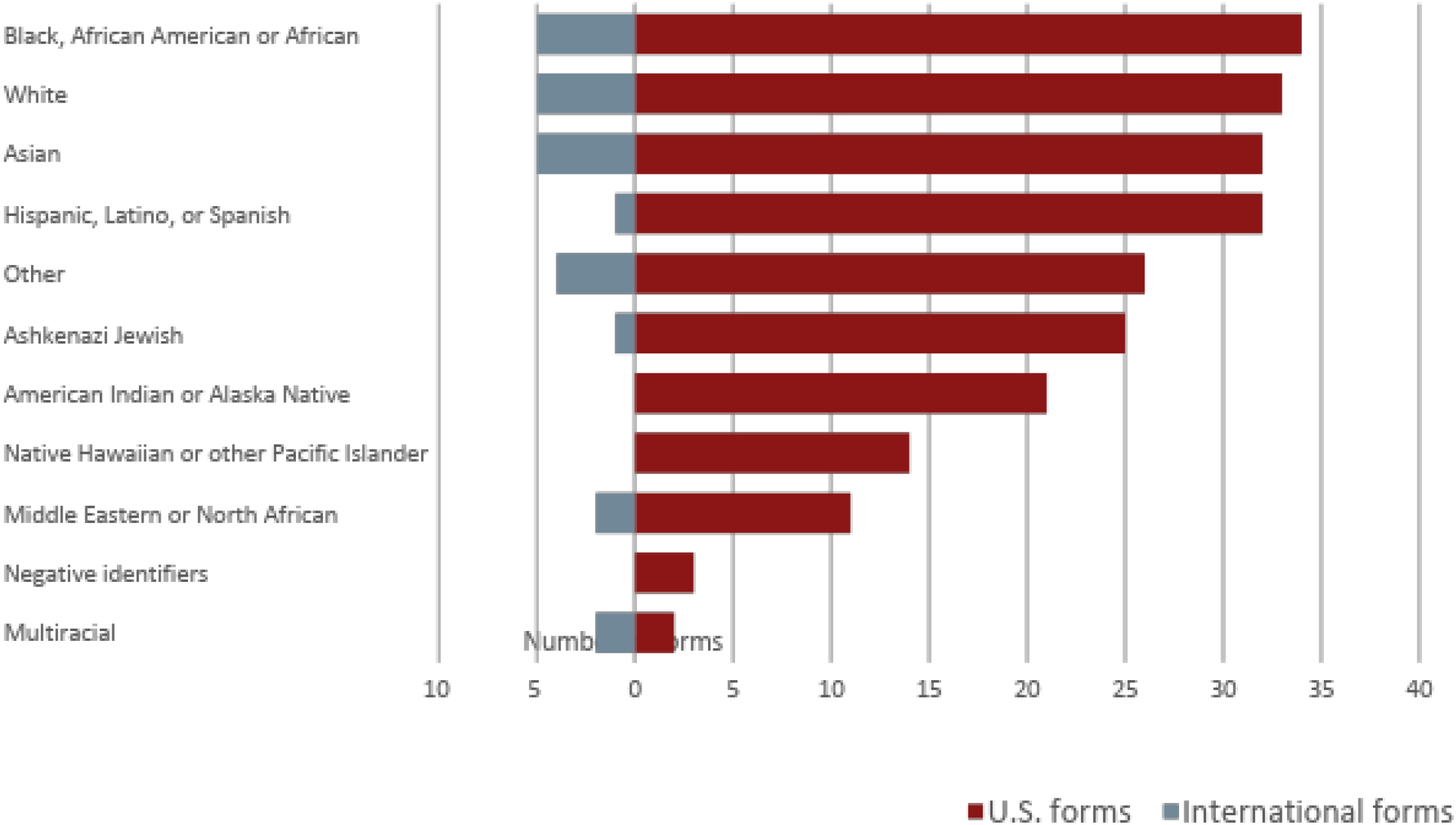
Number of population descriptors (categories) included on RFs from U.S. laboratories (N=34) and non-U.S. laboratories (N=5).

**Figure 5.**
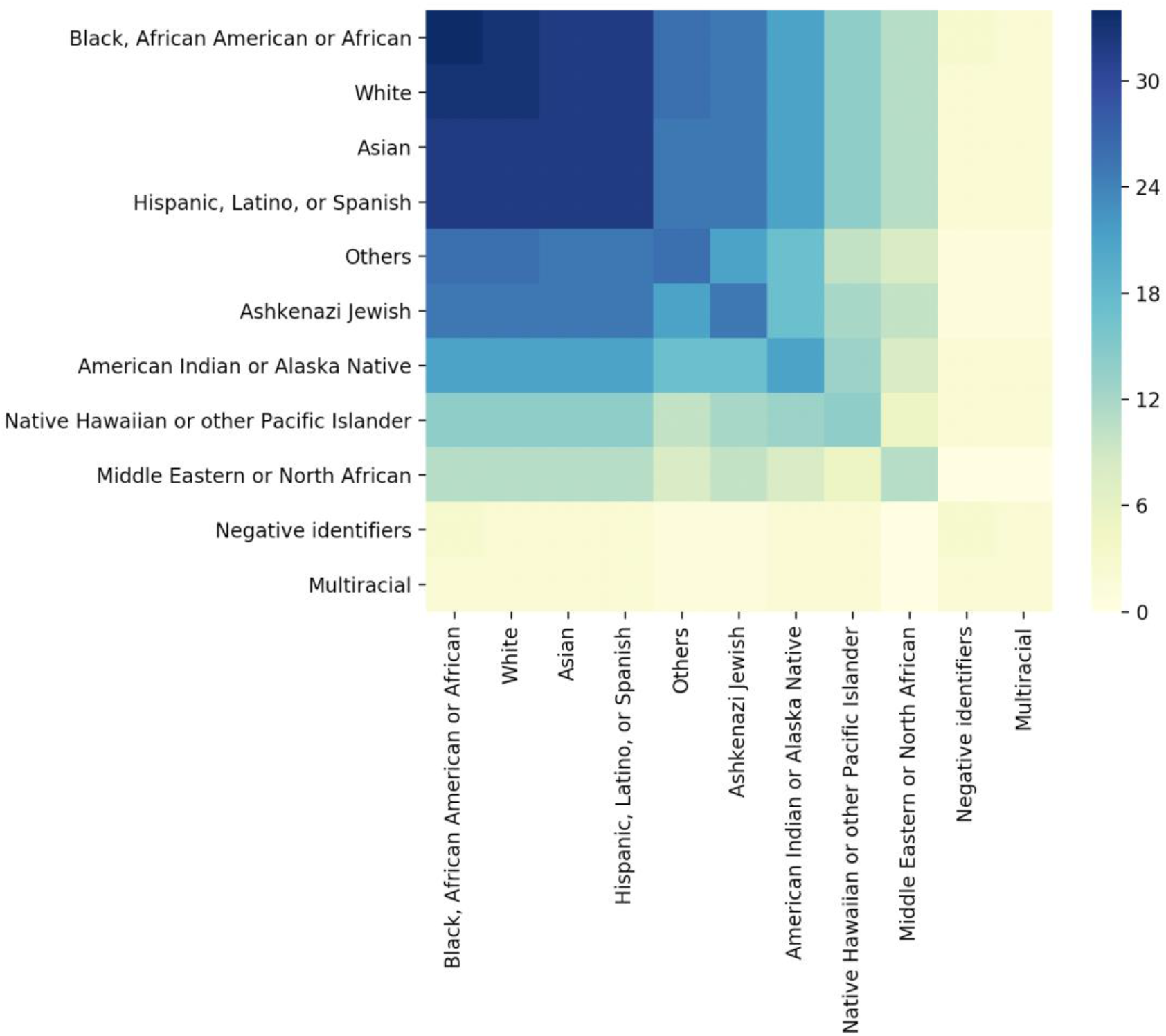
Heat map of co-occurrence of population categories on all RFs analyzed.

There is no clear trend in the relationship between test type and the population framework or descriptors used on clinical lab requisition forms. Those that had no REA input ranged across all test types, from pharmacology to cancer to prenatal/carrier. Within labs, some include more REA categories for prenatal testing compared to other tests, and some include fewer categories.

## DISCUSSION

Information about patient race, ethnicity, and ancestry are more likely to be used in the U.S. context than in other parts of the world. Though this may be due to the vast diversity of people from all different backgrounds in the U.S. compared to some other countries, it may also be the case that historical, socio-cultural, and political forces are responsible for perpetuating the use of such concepts in the U.S. Specifically, ‘race’ is a concept that appears exclusively on RFs from U.S.-based clinical genetics laboratories, whereas other countries’ labs use ‘ethnicity’ or ‘nationality’. There are two central components of our interpretation: 1) the U.S. is more likely to use multiple-choice categorization in clinical genetics because of the pervasiveness of a fixed, discrete paradigm of human categories that have financial and policy implications; and 2) the historical, social, and cultural context of the U.S. perpetuates racial and ethnic classification of individuals into pre-determined groups.

UK and US Census forms provide multiple-choice categories for people to self-identify racially or ethnically. In the UK, these include ‘ethnic groups’: White; Mixed or Multiple; Asian or Asian British; Black, Black British, Caribbean or African; and Other.^11^ The US Office of Management and Budget (OMB) defines seven ‘races’: American Indian or Alaska Native, Asian, Black or African American, Native Hawaiian or Other Pacific Islander, White, and Other; and two ‘ethnicities’: Hispanic and Non-Hispanic.^12^ Some of these broad categories evoke physical attributes such as skin color and facial features, which are associated with socio-cultural stereotypes about racial identity. Others are related to cultural or ethno-linguistic identities, geographic locations, family ancestral origins, nationality, or a combination of these. In each case, concepts of socio-cultural identity and bio-geographic ancestry are conflated or collapsed.

Conceptual inconsistencies obscure the meaning of such broad population categories, even within a single term. For example, UK sub-groups for ‘Asian’ include Indian, Pakistani, Bangladeshi, and Chinese; in the US, they are: Asian Indian, Cambodian, Chinese, Filipino, Hmong, Japanese, Korean, Pakistani, and Vietnamese. There is vast genetic and socio-cultural variation within and among these groupings; in India alone there are >120 languages spoken among >10K people with >2K ethnic groups.^13,14^ As such, the utility of population frameworks that fail to distinguish between Indians, Chinese (>55 ethnic groups^15^), and all others who might be considered racially ‘Asian’ needs to be carefully scrutinized, other than for the purpose of tracking racism.

How meaningful are the designations ‘Asian’, ‘Black’, ‘White’, and ‘Hispanic’ in a research context, or for the purpose of clinical genetic testing? A survey of US clinical genetics professionals revealed that patients’ race and ethnicity are considered important in clinical variant interpretation, and hardly any reported access to genetic ancestry data, despite this being perceived as more relevant.^4^ As such, methods for characterizing patient populations that are currently in use by clinical genetics laboratories need to be examined and revised to maximize utility and minimize harm.

The utilization of race and ethnicity as a proxy for genetic background has the potential to contribute to the furtherance of unsubstantiated beliefs about the genetic underpinnings of race. The impact of environmental factors on health disparities related to societal inequality (i.e., social determinants of health) may be misattributed to genetic factors as a result. As a proxy for historical, social, cultural, and other environmental factors shaped by systemic inequality, the concept of ‘race’ may be an appropriate metric to collect and use in data analyses. However, other measures related to a patient’s environment or lived experience may be preferrable and more readily interpreted. By the same logic, genomic background can be measured or approximated more accurately using family history or other information related to genetics and ancestral origins, as opposed to using race or ethnicity as a proxy variable. Though clinical genetics professionals do not typically have access to genomic data or estimates of genetic ancestry, there may be qualitative approaches to ascertaining ancestry information (i.e., self-report) that could improve inferences, compared to those derived from less relevant (non-genetic) information.

Sampling clinical genetics laboratories that submit the most genetic variants to a U.S.-based resource for clinical variant data sharing, ClinVar. This sampling strategy may have limited our ability to learn about large international labs not included in the analysis, biasing our findings toward the clinical genetics context in the U.S. Clinical laboratories from Germany are also over-represented, relative to other European countries. Language barriers between the investigators and information accessed on clinical genetics laboratory websites (including test requisition forms) may have also prevented accurate identification of population descriptors through translation errors. Finally, some labs may have updated RF demographic questions since the analysis.

Concepts of race, ethnicity and ancestry are commonly encountered in clinical care and biomedical research in the United States. They have the potential to be both empowering and a basis of harmful discrimination. Developing and maintaining an understanding of these concepts from both a present and historical perspective can be helpful in providing culturally appropriate, effective clinical care. Conceptual understanding and knowledge of historical context around race, ethnicity and ancestry enable efforts to ensure that all patients of diverse backgrounds receive optimal care.

## DATA AVAILABILITY

All RFs used in this analysis are publicly available online, and summaries of aggregate data are included in the Supplementary Materials. Additional details are available upon request to the corresponding author.

## ACKNOWLEDGEMENTS

This work was supported in part by Concert Genetics, Inc.; The Clinical Genome Resource (ClinGen; NIH/NHGRI U41HG009649); Stanford University School of Medicine; and the Royal Academy of Engineering of the United Kingdom.

## AUTHOR INFORMATION

Conceptualization: A.B.P, G.H.; Data curation: K.A.; Formal Analysis: A.B.P., J.G.M., K.A.; Funding acquisition: N/A; Investigation: J.G.M; Methodology: A.B.P., J.G.M; Project administration: A.B.P.; Resources: TBD; Software: J.G.M; Supervision: A.B.P., G.H.; Validation: ; Visualization: A.B.P., J.G.M.; Writing – original draft: A.B.P.; Writing – review & editing: A.B.P., J.G.M, K.A., G.H.

## ETHICS DECLARATION

No data from human subjects were included in this research; IRB review not required. The authors report no competing financial interests in relation to the work described.

